# Vascular Proximity Coverage: A Novel CT-Based Marker of Pulmonary Vascular Distance to the Lung Parenchyma

**DOI:** 10.64898/2026.10.15.26365547

**Authors:** Stéphanie Marie Aguilera, Venkata Lakshmi Sthanam, Arie Nakhmani, Joseph M. Reinhardt, Mark T. Dransfield, Surya P. Bhatt, Sandeep Bodduluri

**Affiliations:** Center for Lung Analytics and Imaging Research (CLAIR), University of Alabama at Birmingham, Birmingham, AL 35294; Department of Biomedical Engineering, University of Alabama at Birmingham, Birmingham, AL 35294\; Department of Electrical and Computer Engineering, University of Alabama at Birmingham, Birmingham, AL 35294; Division of Pulmonary, Allergy and Critical Care Medicine, University of Alabama at Birmingham, Birmingham, AL 35294; Roy J. Carver Department of Biomedical Engineering, University of Iowa, Iowa City, IA 52242.

**Keywords:** pulmonary vascular disease, COPD, computed tomography, CT biomarker, vascular proximity coverage

## Abstract

**Rationale:** Conventional CT-derived pulmonary vascular metrics quantify vascular pruning but do not assess spatial heterogeneity of vascular architecture. In COPD, peripheral microvascular pruning coexists with compensatory central vessel dilation, paradoxically preserving total vessel volume while reducing effective vascular penetration into lung parenchyma.

**Objectives:** To develop and validate Vascular Proximity Coverage (VPC), a novel distance-based CT biomarker quantifying spatial efficiency of pulmonary vascular distribution.

**Methods:** This retrospective analysis included 7,881 COPDGene participants with complete vascular segmentations. VPC is estimated by integrating the area under the cumulative lung tissue coverage curve at distances 0–10 mm from segmented vessel voxels. Multivariable models were used to assess associations with lung function, six-minute walk distance (6MWD), St. George’s Respiratory Questionnaire (SGRQ), mMRC dyspnea score and all-cause mortality adjusting for demographics, smoking, emphysema, scanner type, and BV5. We stratified the cohort into four groups defined by the median values of BV5 and VPC, yielding discordant (high-BV5/low-VPC, low-BV5/high-VPC) and concordant (high/high, low/low) vascular phenotypes.

**Results:** VPC declined across disease severity (5.82 ± 0.50 in non-smokers to 5.21 ± 0.64 in GOLD 4; p<0.001). Among spirometrically normal smokers, current smokers showed reduced VPC versus former smokers (5.31 vs. 5.68; p<0.001). Each standard deviation (SD) decrease in VPC was independently associated with reduced 6MWD (β=-16.8 m, 95% CI: -19.5 to -14.0), worse SGRQ (β=1.9, 95% CI: 1.4 to 2.4), and increased dyspnea (RR=1.05, 95% CI: 1.03–1.08; all p<0.001). The lowest VPC quartile, as compared to the reference quartile, was associated with 38% higher mortality (HR=1.38, 95% CI: 1.18–1.62; p<0.001). Based on median thresholds, high-BV5/low-VPC phenotype, compared to High-BV5/High-VPC, showed 24% increased mortality (HR=1.24, 95% CI: 1.08–1.42; p=0.002) despite preserved vessel volume.

**Conclusions:** VPC captures spatial heterogeneity of vascular architecture independent of vessel volume and detects subclinical vascular dysfunction in smokers without airflow obstruction.

**AT A GLANCE COMMENTARY:** *Scientific Knowledge on the Subject:* Current CT-based pulmonary vascular metrics, including volume of blood vessels less than 5 mm² cross-sectional area (BV_5_) and total vessel volume, quantify the amount of visible vasculature but do not assess how effectively vessels reach the lung parenchyma for efficient gas exchange. In COPD, peripheral microvascular pruning commonly coexists with compensatory central vessel dilation, leading to preserved or increased total vascular volume despite compromised functional vascular distribution.

*What This Study Adds to the Field:* Vascular Proximity Coverage (VPC) is a novel distance-based CT biomarker that quantifies the spatial efficiency of pulmonary vascular distribution by measuring cumulative lung tissue coverage across distances from vessel walls. In 7,881 COPDGene participants, including non-smokers and PRISm subjects, lower VPC was independently associated with decreased lung function, reduced exercise capacity, worsened quality of life, and increased all-cause mortality over a median 10.8-year follow-up after adjusting for BV_5_ and emphysema. VPC identified a high-risk phenotype with preserved vessel volume but impaired distribution (high-BV5/low-VPC) that carried 24% increased mortality risk and detected subclinical vascular dysfunction in smokers with normal spirometry.

## INTRODUCTION

Chronic obstructive pulmonary disease (COPD) is a leading cause of morbidity and mortality worldwide (1, 2). While traditionally characterized by airflow limitation, COPD is increasingly recognized as a heterogeneous disease with distinct pathophysiological phenotypes, including those associated with pulmonary vascular abnormalities (3, 4). Pulmonary vascular disease in COPD contributes significantly to exercise limitation, reduced quality of life, and mortality, yet remains undetected due to limitations of current assessment tools (5–8).

Computed tomography (CT) is increasingly being used for non-invasive assessment of pulmonary vascular abnormalities in COPD (7, 8). Current CT-derived vascular metrics include total pulmonary vessel volume and blood volume of vessels with less than 5 mm² cross-sectional area (BV5), a measure thought to reflect small vessel disease (9, 10). These metrics have demonstrated associations with lung function impairment, exercise capacity, and clinical outcomes (11–13). However, these volume-based measures quantify vascular mass without capturing the spatial density of the pulmonary vascular tree, specifically, how effectively vessels distribute throughout the lung parenchyma to facilitate gas exchange (14).

This distinction carries specific relevance in COPD, where vascular pathology often involves heterogeneous changes across the vascular tree. Peripheral vascular pruning, the loss of small vessels in the lung periphery, is a well-documented phenomenon associated with emphysematous destruction (15, 16). Concurrently, central pulmonary vessels may undergo compensatory dilation in response to increased pulmonary vascular resistance (17, 18). This remodeling pattern can paradoxically increase overall visualized vessel volume while simultaneously reducing effective vascular penetration into lung parenchyma, creating a dissociation between volumetric metrics and functional vascular capacity (19).

To address this limitation, we developed the Vascular Proximity Coverage (VPC), a novel distance-based CT biomarker designed to quantify the spatial efficiency of pulmonary vascular distribution. Rather than measuring vascular volume, VPC assesses the proportion of lung parenchyma within functional diffusion distances from vessel walls, directly capturing the concept of vascular proximity. We hypothesized that VPC would provide independent prognostic information beyond existing volumetric metrics and identify high-risk phenotypes characterized by preserved vessel volume but impaired distribution.

## METHODS

### Study Population

This retrospective analysis used data from the COPDGene study, a multicenter observational cohort designed to identify genetic and epidemiological factors associated with COPD (20). The study enrolled 10,198 non-Hispanic White and African American participants aged 45-80 years at 21 clinical centers across the United States between 2008 and 2011. Participants included current and former smokers with at least 10 pack-years of smoking history, as well as non-smoking healthy volunteers. The study protocol was approved by the institutional review boards at all participating centers, and all participants provided written informed consent.

In the current study, we included participants with available inspiratory CT scans. Post-bronchodilator spirometry was performed according to American Thoracic Society recommended guidelines. Participants include non-smokers (never-smokers without airflow obstruction), PRISm (Preserved Ratio with Impaired Spirometry), GOLD 0 (smokers without airflow obstruction, FEV_1_/FVC ≥0.70), and disease severity was assigned according to Global Initiative for Chronic Obstructive Lung Disease (GOLD) spirometric criteria: GOLD 1-4 (21). Six-minute walk distance (6MWD) was measured according to ATS guidelines (22). Health-related quality of life was assessed using the St. George’s Respiratory Questionnaire (SGRQ), with higher scores indicating worse health status (23). Dyspnea was graded using the modified Medical Research Council (mMRC) scale (24). All-cause mortality was ascertained through longitudinal follow-up, supplemented by query of the Social Security Death Index and National Death Index (25). Vital status was determined through December 2024, providing at least 10 years of follow-up from baseline enrollment.

### CT Imaging Protocol

Volumetric CT scans were acquired at full inspiration using multi-detector row CT scanners according to a standardized protocol (20, 26). Scans were performed without intravenous contrast at 120 kVp with tube current modulated based on body habitus. Images were reconstructed using standard algorithms at 0.5-0.75 mm slice thickness. Automated pulmonary vascular segmentation was performed using an in-house UNET algorithm fine-tuned over open-source TotalSegmentator platform (27,28). BV5, defined as the volume of vessels with a cross-sectional area less than 5 mm^2^, and total vessel volume were computed from segmented vascular masks as previously described (8). Emphysema was quantified as the percentage of lung voxels with attenuation less than -950 Hounsfield units on inspiratory CT (%LAA-950) (29).

### Vascular Proximity Coverage (VPC)

VPC was designed to quantify the spatial efficiency of pulmonary vascular distribution by measuring the proportion of lung parenchyma within specified distances from the nearest vessel wall. The calculation proceeds in three steps: First, a Euclidean distance transform is applied to the lung parenchyma mask using the segmented vascular tree as the reference structure. This generates a distance map in which each voxel within the lung is assigned a value representing its minimum distance (in millimeters) to the nearest vessel wall. Second, coverage curves represent the proportion of total lung volume within progressively increasing distance thresholds (from 0.5 to 10 mm in 0.5 mm increments). This generates a coverage distribution curve describing the spatial relationship between vessels and lung parenchyma (**Figure 1, right**). Third, the VPC is calculated by integrating the distribution curve from 0 to 10 mm range using the trapezoidal rule (30). Higher VPC values indicate more efficient vascular distribution, with greater proportions of lung parenchyma near vessels (**Figure 1, left**).

**Figure 1.**
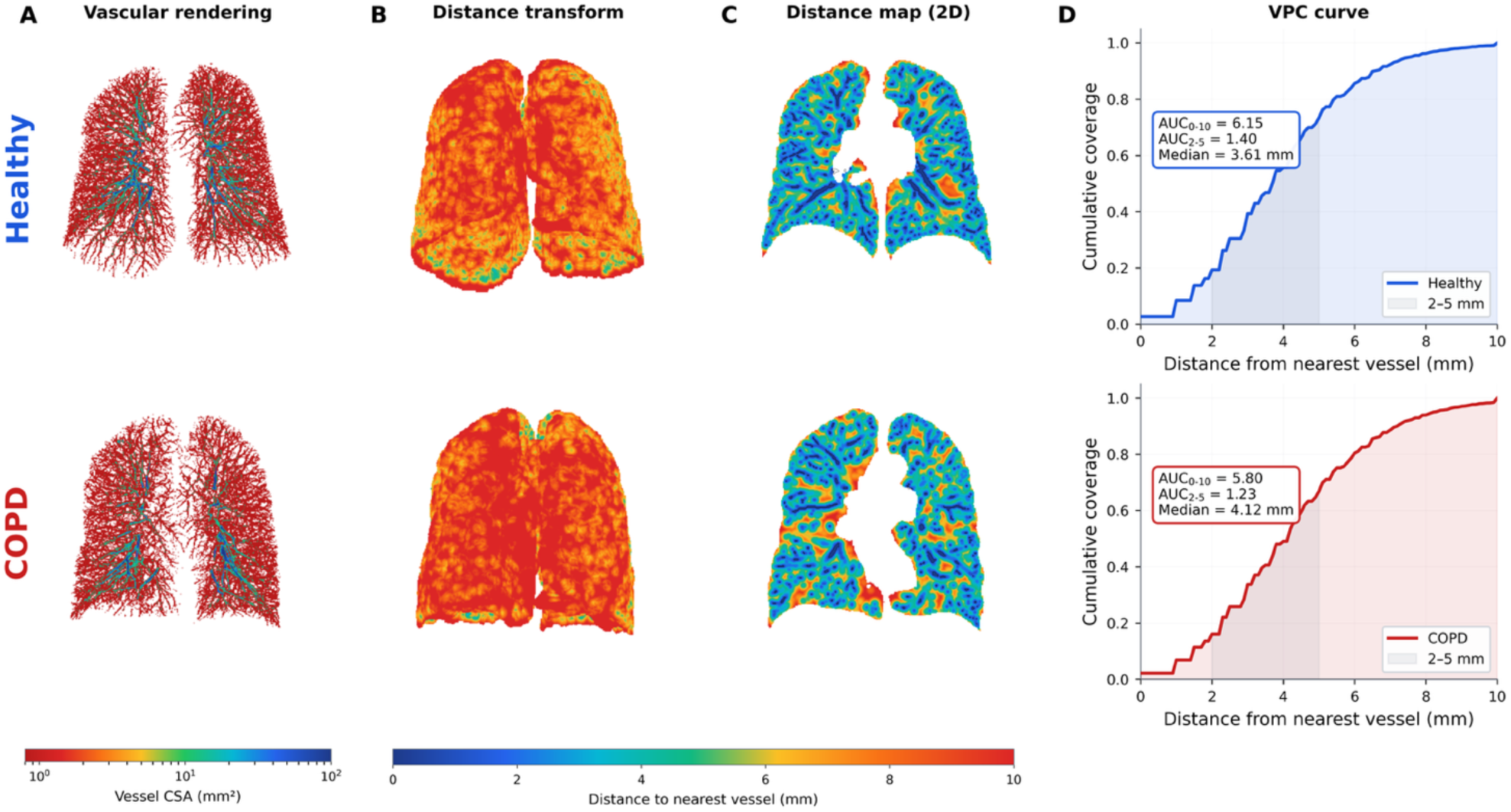
Vascular Proximity Coverage estimation in healthy and COPD subjects.

### Statistical Analysis

Continuous variables are presented as mean ± standard deviation or median (interquartile range) as appropriate; categorical variables are presented as counts (percentages). Differences across GOLD stages were assessed using ANOVA with Tukey post-hoc tests for continuous variables and chi-squared tests for categorical variables. Multivariable linear regression models were used to assess associations between VPC and patient outcomes (FEV_1_, FEV_1_/FVC, 6MWD, SGRQ, mMRC). Models were adjusted for age, sex, race, body mass index (BMI), smoking status, pack-years, scanner type, %LAA-950, and BV5. VPC was standardized (mean=0, SD=1) to facilitate interpretation of effect sizes per standard deviation change. Cox proportional hazards regression, additionally adjusted for FEV1, assessed associations between VPC and all-cause mortality. For quartile analysis, VPC was categorized into quartiles labeled as Normal (highest VPC, best distribution), Mild reduction, Moderate reduction, and Severe reduction (lowest VPC), with the Normal quartile serving as the reference group. Combined phenotype analysis was performed by dichotomizing participants at median values for both BV5 and VPC, creating four groups: high-BV5/high-VPC (reference), high-BV5/low-VPC (preserved volume/impaired distribution), low-BV5/high-VPC (reduced volume/preserved distribution), and low-BV5/low-VPC (combined impairment). All analyses were performed using R version 4.1.0 (R Foundation for Statistical Computing, Vienna, Austria). Two-sided p-values <0.05 were considered statistically significant.

## RESULTS

### Study Population Characteristics

The cohort included 105 non-smokers, 956 PRISm, 3,387 GOLD 0, 603 GOLD 1, 1,508 GOLD 2, 865 GOLD 3, and 457 GOLD 4 participants (**Table 1**). Mean age was 59.5 ± 9.0 years, 53.4% were male, and 66.2% were non-Hispanic White. Mean pack-years among ever-smokers was 43.6 ± 24.3. During median follow-up of 10.8 years (interquartile range [IQR], 5.4–12.9 years), 2,191 deaths occurred (27.8%).

**Table 1.** Baseline Characteristics of the Study Population by GOLD Stage

| Characteristic | Non-smokers (n=105) | PRISm (n=956) | GOLD 0 (n=3,387) | GOLD 1 (n=603) | GOLD 2 (n=1,508) | GOLD 3 (n=865) | GOLD 4 (n=457) |
| --- | --- | --- | --- | --- | --- | --- | --- |
| Age, years | 61.3 ± 9.1 | 57.0 ± 8.1 | 56.4 ± 8.3 | 61.6 ± 8.9 | 62.1 ± 8.9 | 64.1 ± 8.3 | 64.0 ± 7.5 |
| Male, n (%) | 33 (40) | 447 (44) | 1804 (45) | 340 (45) | 808 (46) | 506 (45) | 276 (46) |
| African American, n (%) | 7 (8) | 428 (45) | 1409 (42) | 142 (27) | 376 (28) | 179 (21) | 86 (19) |
| BMI, kg/m<sup>2</sup> | 28.0 ± 5.0 | 32.0 ± 7.2 | 29.0 ± 5.8 | 27.1 ± 4.9 | 28.8 ± 6.1 | 28.0 ± 6.3 | 25.3 ± 5.5 |
| Pack-years | - | 41.8 ± 23.1 | 37.3 ± 20.2 | 44.8 ± 24.3 | 50.5 ± 26.8 | 54.9 ± 26.4 | 56.3 ± 28.9 |
| Current smoker, % | - | 613 (46) | 2061 (44) | 347 (44) | 755 (44) | 327 (41) | 107 (26) |
| FEV<sub>1</sub>, L | 2.90 ± 0.70 | 2.07 ± 0.50 | 2.90 ± 0.67 | 2.66 ± 0.67 | 1.89 ± 0.51 | 1.16 ± 0.30 | 0.65 ± 0.19 |
| FEV<sub>1</sub>/FVC | 0.80 ± 0.05 | 0.77 ± 0.05 | 0.79 ± 0.05 | 0.65 ± 0.04 | 0.59 ± 0.08 | 0.44 ± 0.09 | 0.31 ± 0.07 |
| %LAA-950 | 2.0 ± 2.7 | 1.4 ± 2.6 | 1.9 ± 2.5 | 5.1 ± 5.7 | 7.0 ± 7.8 | 16.0 ± 12.7 | 26.7 ± 13.9 |
| 6MWD, m | 518 ± 99 | 387 ± 110 | 454 ± 106 | 457 ± 103 | 397 ± 111 | 336 ± 113 | 269 ± 107 |
| SGRQ total score | 2.9 ± 6.0 | 29.7 ± 23.1 | 16.9 ± 17.9 | 18.5 ± 17.9 | 33.2 ± 21.7 | 46.3 ± 19.3 | 55.6 ± 16.6 |
| mMRC dyspnea score | 0.1 ± 0.4 | 1.5 ± 1.5 | 0.8 ± 1.2 | 0.8 ± 1.2 | 1.6 ± 1.4 | 2.5 ± 1.2 | 3.1 ± 0.9 |
| VPC | 5.82 ± 0.50 | 5.43 ± 0.76 | 5.45 ± 0.66 | 5.42 ± 0.57 | 5.41 ± 0.68 | 5.30 ± 0.67 | 5.21 ± 0.64 |
| BV5, mL | 164.6 ± 34.9 | 145.5 ± 38.2 | 160.6 ± 36.9 | 177.1 ± 40.8 | 165.6 ± 36.5 | 160.5 ± 35.2 | 158.1 ± 32.3 |

Data presented as mean ± SD, median (IQR), or n (%). NHW = non-Hispanic White; BMI = body mass index; FEV<sub>1</sub> = forced expiratory volume in 1 second; FVC = forced vital capacity; %LAA-950 = percentage of low-attenuation areas below -950 HU; 6MWD = six-minute walk distance; SGRQ = St. George's Respiratory Questionnaire; mMRC = modified Medical Research Council dyspnea scale; VPC = Vascular Proximity Coverage area under curve 0–10 mm. PRISm = Preserved Ratio Impaired Spirometry.

### VPC Distribution Across Disease Severity

VPC decreased across disease severity categories (**Figure 2**). Non-smokers had the highest VPC (5.82 ± 0.50). VPC was lower in smokers without airflow obstruction (5.45 ± 0.66) compared to non-smokers (p<0.001), PRISm participants showed reduced VPC (5.43 ± 0.76), similar to smokers without airflow obstruction. VPC decreased progressively from GOLD 1 (5.42 ± 0.57) through GOLD 2 (5.41 ± 0.68), GOLD 3 (5.30 ± 0.67), to GOLD 4 (5.21 ± 0.64; Jonckheere-Terpstra p<0.001 for trend across GOLD 0–4). Within the GOLD 0 population, current smokers demonstrated significantly lower VPC compared to former smokers (5.31 ± 0.68 vs. 5.68 ± 0.56, p<0.001).

**Figure 2.**
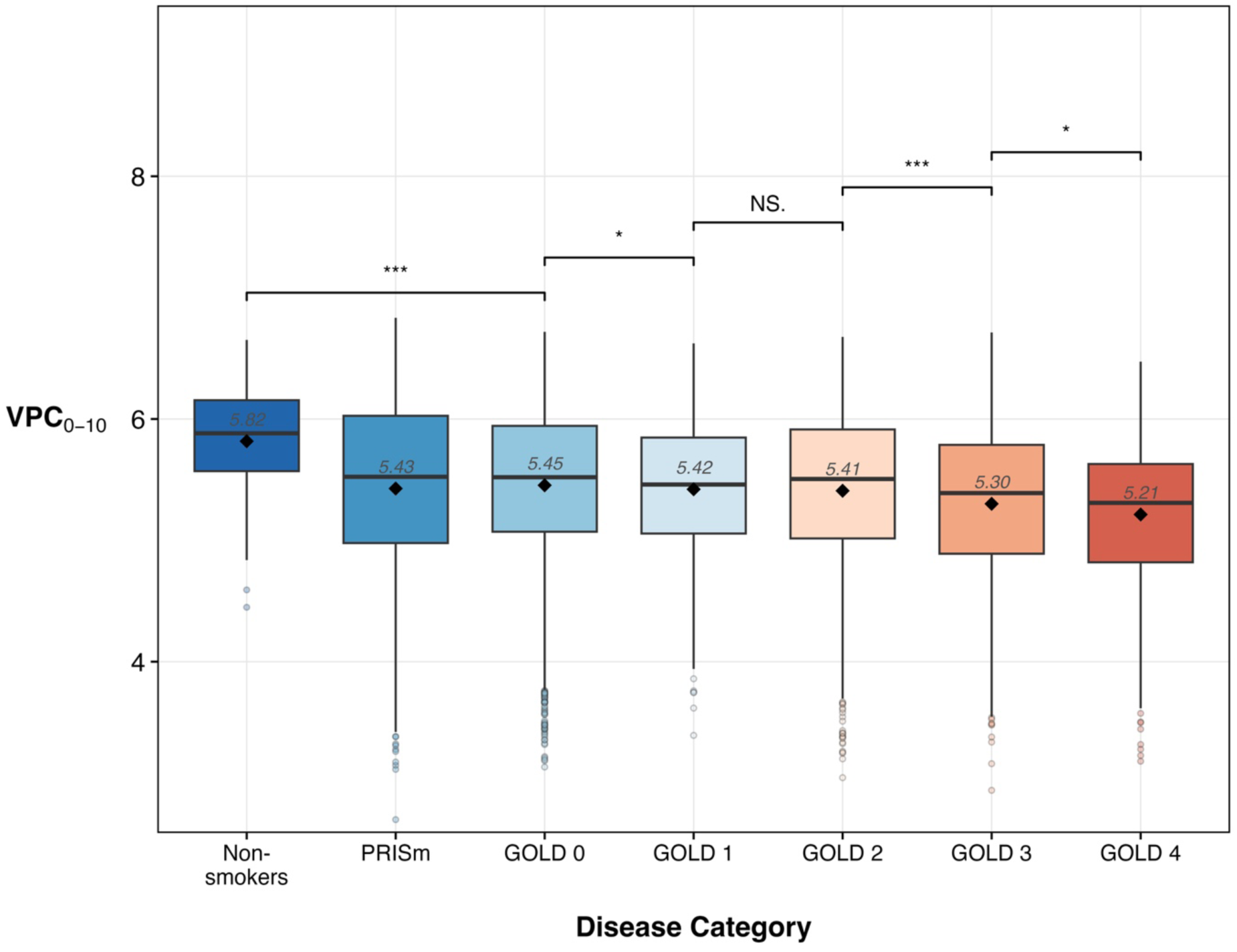
Vascular Proximity Coverage (VPC) by GOLD Stage. Box plots showing VPC distribution across disease severity categories, including non-smokers, PRISm, and GOLD 0–4. VPC distributions are shown for non-smokers, PRISm, and GOLD 0–4. Note that PRISm is shown separately as it is not on the GOLD severity continuum. VPC is highest in non-smokers (5.82 ± 0.50) and lowest in GOLD 4 (5.21 ± 0.64). Significance brackets indicate pairwise comparisons: *p<0.05, **p<0.01, ***p<0.001. Diamond markers show group means; boxes represent interquartile range with median line; whiskers extend to 1.5× IQR.

### Relationship Between VPC and BV5

VPC demonstrated a weak correlation with BV5 (Pearson r=0.11, p<0.001) (**Figure 3**). Scatter plot analysis revealed substantial heterogeneity in VPC at any given BV5 level.

**Figure 3.**
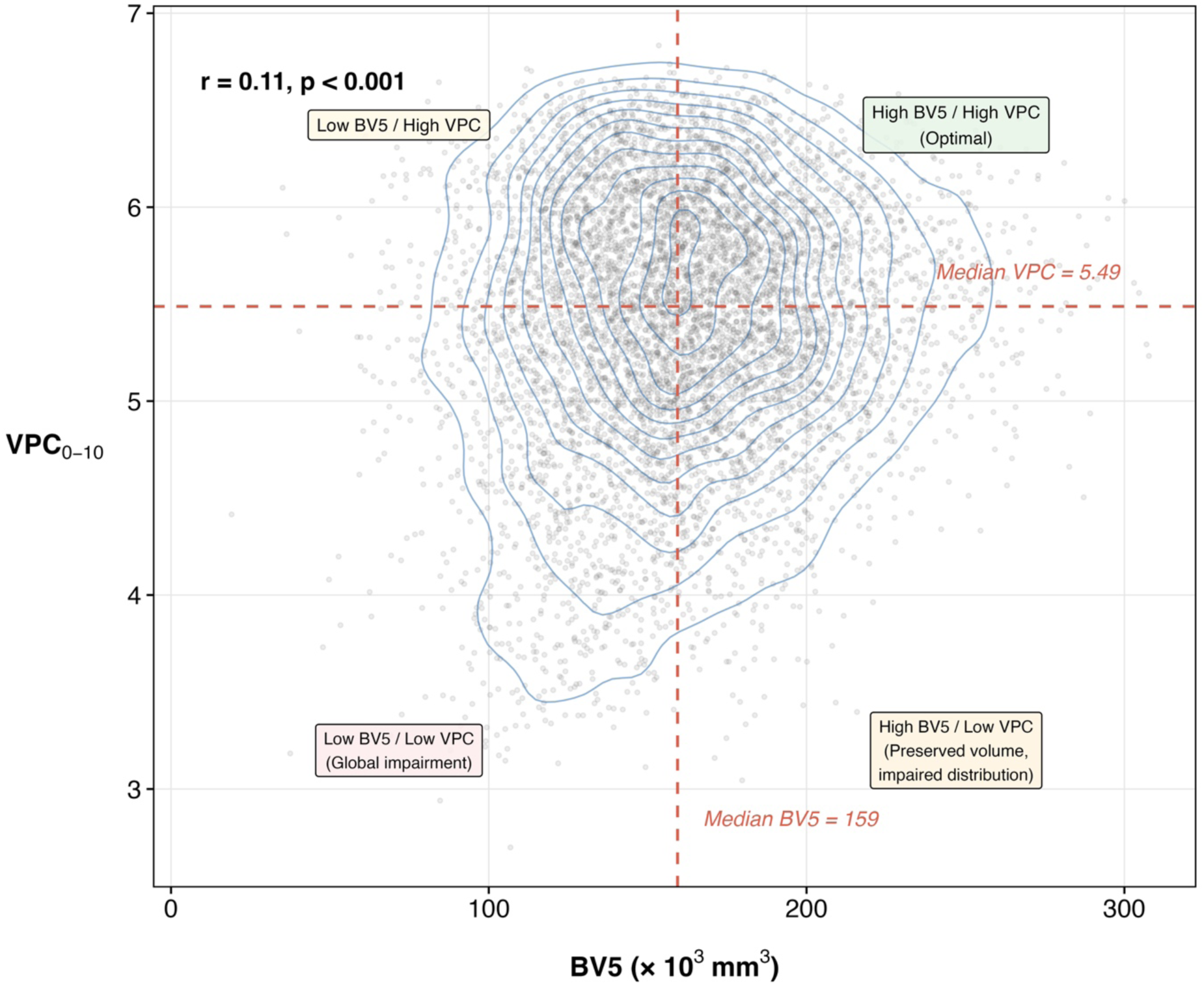
Relationship between VPC and BV5. The scatter plot demonstrates a weak correlation between VPC and BV5 (Pearson r=0.11, p<0.001). Dashed lines indicate median values used to define combined phenotype groups. Density contours illustrate the joint distribution. Substantial heterogeneity in VPC at any given BV5 level indicates these metrics capture independent aspects of vascular structure.

### Associations with Clinical Outcomes

In multivariable models adjusted for demographics, smoking, emphysema, scanner type, and BV_5_, VPC demonstrated significant independent associations with multiple clinical outcomes (**Table 2**).

**Table 2.** Multivariable Associations Between VPC and Clinical Outcomes

| Outcome | $\beta$ or HR | 95% CI | p-value | Model |
| --- | --- | --- | --- | --- |
| **Lung Function** |  |  |  |  |
| FEV<sub>1</sub>, L | $\beta = -0.055$ | -0.081, -0.028 | <0.001 | GLM |
| FEV<sub>1</sub>/FVC | $\beta = -0.0066$ | -0.0095, -0.0037 | <0.001 | GLM |
| **Functional Capacity & Symptoms** |  |  |  |  |
| 6MWD, m | $\beta = -16.75$ | -19.48, -14.01 | <0.001 | GLM |
| SGRQ total score | $\beta = 1.90$ | 1.39, 2.42 | <0.001 | GLM |
| mMRC dyspnea score | RR = 1.05 | 1.03, 1.08 | <0.001 | Poisson |
| **All-Cause Mortality (VPC Quartiles)<sup>†</sup>** |  |  |  |  |
| Normal (high-VPC) | Reference |  |  | Cox PH |
| Mild reduction | HR = 1.20 | 1.04, 1.38 | 0.013 | Cox PH |
| Moderate reduction | HR = 1.22 | 1.05, 1.42 | 0.008 | Cox PH |
| Severe reduction | HR = 1.38 | 1.18, 1.62 | <0.001 | Cox PH |
| **All-Cause Mortality (BV5-VPC Phenotypes)<sup>‡</sup>** |  |  |  |  |
| High-BV5 / High-VPC (optimal) | Reference |  |  | Cox PH |
| High-BV5 / Low-VPC | HR = 1.24 | 1.08, 1.42 | 0.002 | Cox PH |
| Low-BV5 / High-VPC | HR = 1.20 | 1.03, 1.38 | 0.016 | Cox PH |
| Low-BV5 / Low-VPC (global) | HR = 1.32 | 1.14, 1.52 | <0.001 | Cox PH |

*All models adjusted for age, sex, race, BMI, smoking status, pack-years, scanner type, %LAA-950, and BV5 (except phenotype model). VPC standardized (mean=0, SD=1) for continuous analyses. Mortality models additionally adjusted for FEV<sub>1</sub> for regression and quartile models (complete cases); $\beta$ = regression coefficient per 1 SD decrease in VPC; HR = hazard ratio; RR = rate ratio; CI = confidence interval; GLM = generalized linear model; Cox PH = Cox proportional hazards. <sup>†</sup>Reference group is the highest VPC quartile (Normal, best vascular distribution). <sup>‡</sup>Phenotype model adjusts for all covariates except BV5 (used in phenotype definition). C-index = 0.73 (quartile model) and 0.73 (phenotype model).*

### Lung Function

Each standard deviation decrease in VPC was associated with 55 mL lower FEV_1_ (95% CI: 28–81 mL, p<0.001). VPC was also independently associated with lower FEV_1_/FVC ratio (β=-0.007, 95% CI: -0.010 to -0.004, p<0.001) after adjustment for covariates including BV_5_ and emphysema.

### Functional Capacity

Lower VPC was associated with reduced 6MWD (β=-16.8 m per SD, 95% CI: -19.5 to -14.0 m, p<0.001). This association persisted even after adjustment for FEV_1_ and emphysema.

### Respiratory Symptoms

After adjusting for covariates, each standard deviation decrease in VPC was associated with 1.9-point higher SGRQ total score (β=1.90, 95% CI: 1.39 to 2.42, p<0.001) and higher mMRC dyspnea score (RR=1.05, 95% CI: 1.03–1.08, p<0.001).

### Mortality Analysis

In quartile analysis (n=1,983 per quartile), compared to the highest VPC quartile, hazard ratios progressively increased: mild quartile (HR=1.20, 95% CI: 1.04–1.38, p=0.013), moderate reduction quartile (HR=1.22, 95% CI: 1.05–1.42, p=0.008), and severe reduction quartile (HR=1.38, 95% CI: 1.18–1.62, p<0.001) after adjustment for age, sex, race, BMI, smoking status, pack-years, FEV_1_, scanner type, emphysema, and BV5 (**Figure 4**). The model demonstrated good discrimination (C-index=0.739 for the quartile model).

**Figure 4.**
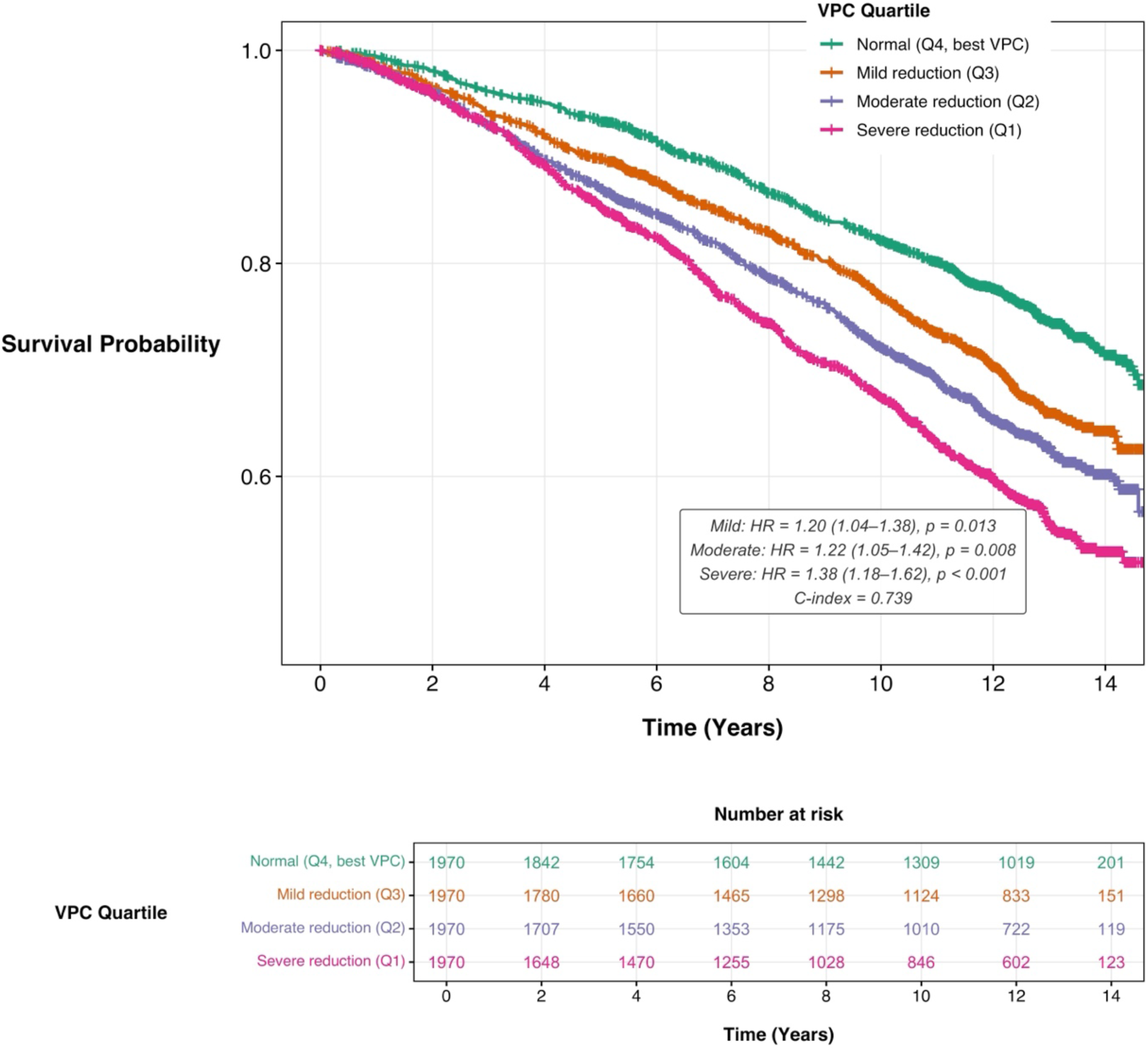
Kaplan-Meier Survival Curves by VPC Quartile. All-cause mortality stratified by VPC quartile with number-at-risk table. Progressive separation of curves demonstrates a graded relationship between vascular distribution efficiency and all-cause mortality over a median follow-up of 10.8 years. Compared to the best quartile (Normal), the severe quartile showed 38% increased mortality (HR=1.38, 95% CI: 1.18–1.62, p<0.001). C-index=0.742. Quartile boundaries: Normal (VPC ≥5.91), Mild (5.49–5.91), Moderate (5.02–5.49), Severe (VPC <5.02).

### Multivariable Combined BV5-VPC Phenotype Analysis

Stratifying participants by median BV5 (159,404 mm³) and median VPC (5.49) values revealed distinct risk profiles (**Figure 5**). Using the reference phenotype (high-BV5/high-VPC; n=1,705) as reference, low-BV5/high-VPC phenotype (n=1,605) showed 20% increased mortality risk (HR=1.20, 95% CI: 1.03–1.38, p=0.016). The high-BV5/low-VPC phenotype (n=1,587), representing preserved vessel volume but impaired spatial distribution, had 24% increased mortality (HR=1.24, 95% CI: 1.08–1.42, p=0.002), while the combined impairment phenotype (low-BV5/low-VPC; n=1,688) demonstrated the highest mortality (HR=1.32, 95% CI: 1.14–1.52, p<0.001)

**Figure 5.**
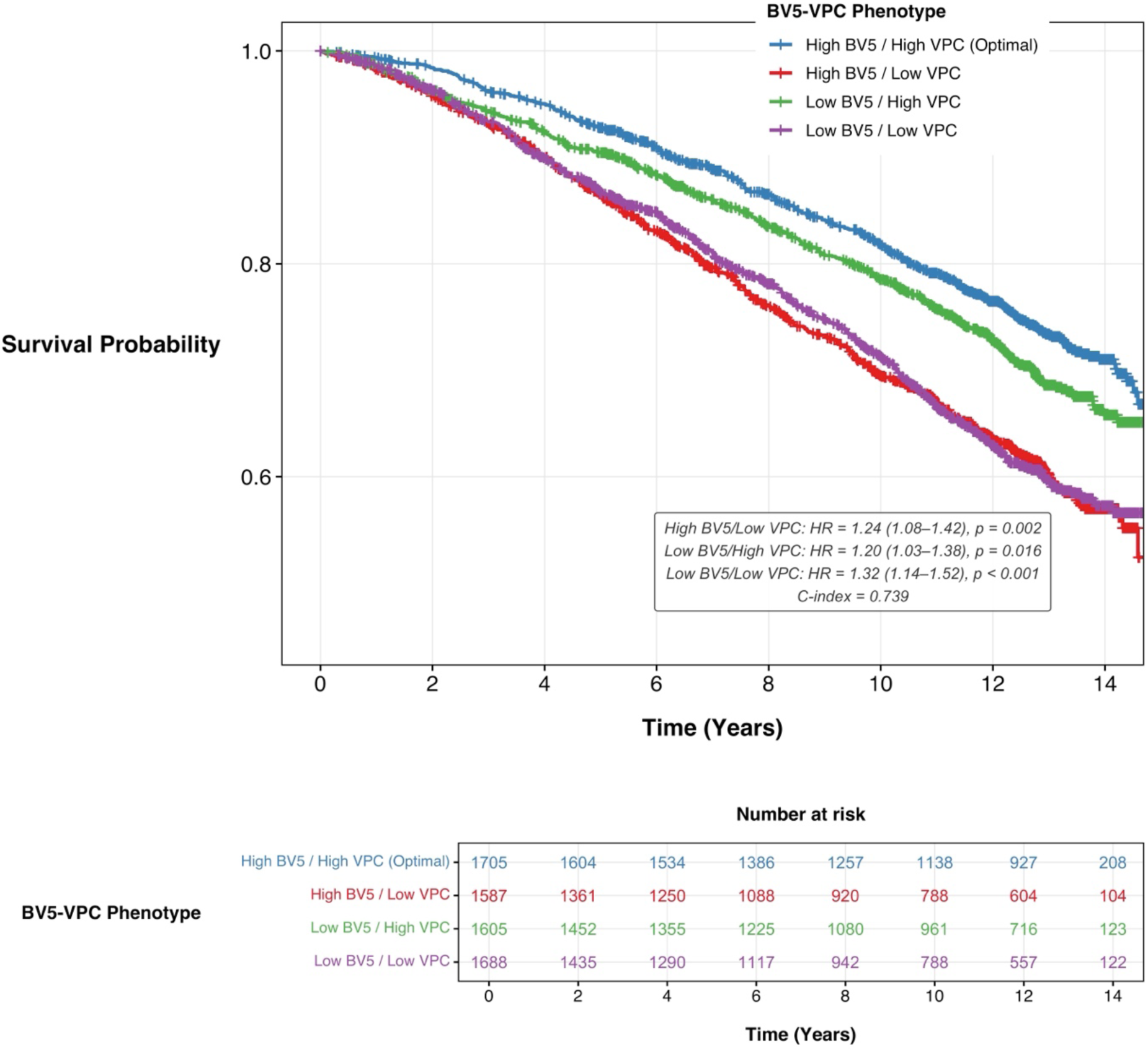
Combined BV5-VPC Phenotype Survival Analysis. Kaplan-Meier curves for four phenotypes defined by median BV5 (159,404 mm³) and median VPC (5.49) with the number-at-risk table. High-BV5/high-VPC (reference n=1,705) serves as reference. High-BV5/low-VPC (preserved volume/impaired distribution; n=1,587) shows 24% increased mortality (HR=1.24, p=0.002) despite preserved vessel volume. Low-BV5/high-VPC (n=1,605) shows 20% increased risk (HR=1.20, p=0.016). Low-BV5/low-VPC (combined impairment; n=1,688) demonstrates the highest mortality (HR=1.32, p<0.001). C-index=0.739.

## DISCUSSION

The Vascular Proximity Coverage (VPC) is a novel CT-based biomarker that quantifies the spatial efficiency of pulmonary vascular distribution. Our principal findings are: (1) lower VPC is independently associated with reduced functional capacity, worse respiratory quality of life, and increased respiratory symptoms after adjustment for conventional vascular metrics and emphysema; (2) VPC predicts all-cause mortality with a graded dose-response relationship; (3) VPC identifies a clinically important high-risk phenotype with preserved vessel volume but impaired distribution; and (4) VPC detects subclinical vascular dysfunction in smokers with normal spirometry.

The conceptual framework underlying VPC addresses a fundamental limitation of existing CT vascular metrics. Current measures such as BV5 and total vessel volume quantify global vascular density but cannot distinguish between efficient and inefficient spatial distributions. In healthy lungs, the pulmonary vascular tree exhibits fractal branching patterns that optimize coverage of the lung parenchyma for gas exchange (25, 31). COPD disrupts this architecture through heterogeneous mechanisms: peripheral vascular pruning reduces small vessel coverage while central vessels may dilate in response to increased pulmonary vascular resistance (15–18). These countervailing changes can maintain or even increase total visualized vessel volume while substantially reducing effective vascular reach.

The weak correlation between VPC and BV5 suggests that these metrics capture distinct aspects of vascular structure. The substantial heterogeneity in VPC at any given BV5 level shows the independent information provided by each metric. This has important clinical implications: patients with similar BV5 values may have markedly different vascular distribution patterns and, consequently, different functional capacities and prognoses.

The combined phenotype analysis provides mechanistic insight into distinct patterns of pulmonary vascular disease. The high-BV5/low-VPC phenotype carried 24% increased mortality despite apparently normal volumetric metrics, likely reflecting central vessel dilation compensating for peripheral vascular loss. Histopathological studies have documented both peripheral vascular loss (rarefaction) and proximal vessel remodeling in COPD (32, 33), though their relative contributions to clinical outcomes have been difficult to disentangle. VPC provides a non-invasive means to characterize these distinct patterns of vascular remodeling. The worst outcome was observed in participants with combined vascular impairment (low-BV5/low-VPC; HR=1.32), consistent with the combination of pruning and poor spatial distribution representing the most advanced vascular pathology.

A major strength of VPC is its ability to detect subclinical vascular dysfunction in smokers without airflow obstruction. Current smokers without airflow obstruction demonstrated significantly reduced VPC compared to former smokers (5.31 vs. 5.68, p<0.001), indicating that vascular architectural changes precede measurable airflow limitation. This finding aligns with pathological evidence that pulmonary vascular remodeling occurs early in smoking-related lung disease (34, 35). VPC may therefore have utility for identifying individuals at risk for poor outcomes before conventional diagnostic criteria are met. The inclusion of PRISm participants adds another dimension to vascular phenotyping in COPD. PRISm subjects showed VPC values (5.43 ± 0.76) comparable to GOLD 1 (5.42 ± 0.57), suggesting similar degrees of vascular maldistribution despite different spirometric profiles. Given the established poor outcomes in PRISm (36), VPC may help elucidate the vascular contributions to morbidity in this understudied population. The independent associations between VPC and clinical outcomes after adjustment for BV5 and emphysema suggest that vascular distribution could contribute to COPD morbidity through mechanisms beyond airway and parenchymal disease, including impaired gas exchange efficiency, ventilation-perfusion mismatch, and reduced pulmonary vascular reserve during exercise (37, 38).

Our analysis has several limitations. First, our segmentation did not distinguish pulmonary arteries from veins, and VPC reflects the combined distribution. While arterial distribution is most relevant for gas exchange, combined arteriovenous assessment may provide a reasonable approximation of overall vascular coverage. Second, VPC quantifies vascular proximity but cannot assess perfusion directly; integration with dual-energy CT perfusion imaging or functional MRI could address this limitation. Third, COPDGene participants are predominantly current or former smokers, and generalizability to never-smoking COPD phenotypes requires confirmation. Fourth, while we adjusted for scanner manufacturer, residual technical variation across the 21 clinical centers may influence VPC measurements; future studies should assess test-retest reproducibility.

In conclusion, VPC represents a novel approach to CT-based vascular assessment that captures functional architecture independent of vessel volume. By quantifying the spatial efficiency of pulmonary vascular distribution, VPC identifies high-risk phenotypes missed by volumetric metrics alone and detects subclinical dysfunction in smokers without airflow obstruction. This metric may enable risk stratification and guide patient selection for trials of vascular-targeted therapies in COPD.

## Data Availability

COPDGene data are available to qualified investigators through the COPDGene Study (www.copdgene.org) under its established data-access procedures.

## Author Contributions

*Study concept and design*: Aguilera, Bhatt, and Bodduluri

*Acquisition, analysis, or interpretation of data*: All authors

*Drafting of the manuscript*: Aguilera, Bhatt, and Bodduluri

*Critical revision of the manuscript for important intellectual content*: All authors

*Statistical analysis*: Aguilera and Bodduluri

*Study supervision*: All authors

